# The association of lifestyle with cardiovascular and all-cause mortality based on machine learning: A Prospective Study from the NHANES

**DOI:** 10.1101/2024.02.07.24302473

**Authors:** Xinghong Guo, Jian Wu, Mingze Ma, Clifford Silver Tarimo, Fengyi Fei, Lipei Zhao, Beizhu Ye

## Abstract

**Objective:** To develop a machine learning (ML) risk stratification model for predicting all-cause mortality and cardiovascular mortality while estimating the influence of lifestyle behavioral factors on the model’s efficacy.

**Method:** A prospective cohort study was conducted using a nationally representative sample of adults aged 40 years or older, drawn from the US National Health and Nutrition Examination Survey from 2007 to 2010. The participants underwent a comprehensive in-person interview and medical laboratory examinations, and subsequently, their records were linked with the National Death Index for further analysis.

**Result:** Within a cohort comprising 7921 participants, spanning an average follow-up duration of 9.75 years, a total of 1911 deaths, including 585 cardiovascular-related deaths, were recorded. The model predicted mortality with an area under the receiver operating characteristic curve (AUC) of 0.848 and 0.829. Stratifying participants into distinct risk groups based on ML scores proved effective. All lifestyle behaviors exhibited an inverse association with all-cause and cardiovascular mortality. As age increases, the discernible impacts of dietary scores and sedentary time become increasingly apparent, whereas an opposite trend was observed for physical activity.

**Conclusion:** We develop a ML model based on lifestyle behaviors to predict all-cause and cardiovascular mortality. The developed model offers valuable insights for the assessment of individual lifestyle-related risks. It applies to individuals, healthcare professionals, and policymakers to make informed decisions.

## Introduction

Cardiovascular disease (CVD) poses a formidable challenge to global health, contributing significantly to non-communicable diseases (NCDs) and representing a leading cause of mortality worldwide^1,2^. According to data from the World Health Organization (WHO), cardiovascular disease contributes to deaths of nearly one million people in the United States, accounting for 30% of the total annual mortality^3^. The escalating prevalence of CVD over the past few decades underscores the urgency of identifying effective preventive measures. Extensive research has elucidated a link between modifiable lifestyles and cardiovascular mortality.^4–7^ The inherent modifiability of lifestyle renders it of considerable practical significance as a predictive model factor. Through model prediction, the population can be informed of the current level of risk in their lifestyle and effectively promote their transition to a healthier lifestyle. However, traditional statistical methods exhibit limitations in establishing predictive models, struggling to effectively handle the intricate interaction between numerous variables.

Machine learning (ML), with its ability to analyze vast and complex datasets, presents a compelling solution to the limitations of traditional methods in unraveling the multifaceted associations between lifestyle choices and mortality outcomes^8^. Unlike conventional statistical models that rely on predefined hypotheses and assumptions, ML algorithms can identify intricate patterns and nonlinear relationships within data, offering a more holistic and data-driven perspective^9,10^. In recent times, an increasing number of studies have applied ML in the field of cardiovascular disease.^11,12^. This becomes particularly crucial in the realm of cardiovascular health, where the impact of diverse lifestyle factors may manifest in subtle and interconnected manners.

The NHANES dataset holds a distinct advantage due to its comprehensive inclusion of health, lifestyle, and biochemical information, providing a rich data source for analysis^13–15^. Implementing of high-quality standardized collection and testing procedures effectively mitigates the potential for measurement bias, ensuring the reliability of the data. This robust data quality, coupled with a wealth of information, facilitates in-depth exploration of the intricate relationship between lifestyle and both cardiovascular and all-cause mortality, offering a reliable and comprehensive foundation for unraveling the complexities inherent in this association.

This study endeavors to establish a predictive model for mortality related to lifestyle factors and aims to delve into the intricate role of these lifestyle factors using ML models.

## Method

The prospective cohort were derived from the National Health and Nutrition Examination Survey (NHANES), a nationwide survey conducted biennially since 1999. All NHANES protocols received approval from the National Center for Health Statistics ethics review board, and written informed consent was obtained from all participants. The modeling survey was deemed exempt from further review.

### Study Population

The sample population was derived from the NHANES cycles of 2007-2008 and 2009-2010. We selected participants aged over 40 who participated in in-person interview, physical examinations and laboratory tests in a mobile examination center. The screening process is shown in **Supplementary Figure 1**.

### Study outcomes

The follow-up data was obtained from the National Health Data Center, which links the NHANES survey population with the death records of the National Death Index (NDI).

Cardiovascular mortality was determined using the International Statistical Classification of Diseases, 10th Revision (ICD-10), and the NCHS classified cardiovascular diseases (054-068, 070). We linked participants with the 2019 mortality data records and excluded individuals whose follow-up years and survival status could not be ascertained.

### Model features

The model encompassed a set of features including age, gender, race, BMI, education level, income, hypertension, diabetes, family history of diseases, non HDL-cholesterol, C-reactive protein, diet score, physical activity level, Sedentary minutes, sleep quality, alcohol consumption and smoking status. Age, gender, race, education level, income, and history of close family diseases can be directly obtained from interview data. BMI was derived from physical examination data, while non-HDL cholesterol and C-reactive protein values were obtained from laboratory test data. Sedentary minutes were acquired through the physical activity questionnaire.

NHANES contains a wealth of nutrition information gathered through health interviews, health examinations, and laboratory testing. Participants underwent a 24-hour dietary recall (First Day) interview as part of their health examination at the mobile examination center. Subsequently, they were instructed to complete a second 24-hour dietary recall (Second Day) interview within a period of 3 to 10 days following the initial recall. To rate the dietary patterns of participants, the following steps were taken: linking to the Food Patterns Equivalents Database (FPED) of the US Department of Agriculture based on the USDA code of the food, estimating the daily nutritional intake of participants based on the 24-hour dietary recall on the first day and the 24-hour dietary recall on the second day, and referencing the US Dietary Guidelines 2020-2025 and the scoring rules of the Healthy Eating Index (HEI) to assess and rate the dietary patterns of participants.

Physical activity was obtained from NHANES’s physical activity questionnaire. The questionnaire contains the information on the weekly exercise intensity and corresponding time reported by the participants. Participants were classified into four (4) groups based on the 2^nd^ edition of the Physical Activity Guidelines for Americans. The "Inactive" group comprised individuals not involved in any moderate- or vigorous-intensity physical activity beyond basic daily life movements. Those deemed "Insufficiently active" engaged in some moderate- or vigorous-intensity physical activity but did not reach the threshold of 150 minutes of moderate-intensity activity per week, or 75 minutes of vigorous-intensity activity, or the equivalent combination. The "Active" category encompassed participants achieving the equivalent of 150 to 300 minutes of moderate-intensity physical activity weekly, meeting the key guideline target range for adults. Lastly, the "Highly active" group included individuals undertaking more than 300 minutes of moderate-intensity physical activity weekly, surpassing the key guideline target range for adults.

Due to the J-shaped association between sleep duration and all-cause mortality, participants were divided into three groups based on sleep duration: optimal (6-8 hours/day), intermediate (5-5.9 or 8.1-10 hours/day), and poor (<5 or >10 hours/day)^6,16^.

Smoking status was categorized into three groups: non-smokers, individuals who smoked previously, and those who reported current smoking based on responses to the cigarette use questionnaire. Data on alcohol consumption was derived from alcohol use questionnaire, wherein participants provided information on the frequency and quantity of drinks consumed. The average daily alcohol consumption was used to measure the level of alcohol consumption among participants.

Variables related to mental health exhibiting missing values exceeding 40% were excluded from the analysis. Subsequently, the random forest (RF) algorithm was employed to impute missing values in the remaining dataset. In order to mitigate the influence of dimensionality and enhance modeling efficiency, continuous variables were rescaled and standardized. The data distribution before imputation is presented in **Supplementary Table 1 & 2**.

### Model development and Risk stratification

A binary classification model was constructed based on follow-up data and participant features to predict mortality. Model development included trials of various ML classifiers, including logistic regression, ridge regression, support vector machines, random forest and Extreme Gradient Boosting (XGBoost). The initial step involved cross validation on the selected models to determine the approximate range of optimal values for each parameter followed by deployment of the grid search method to select the best model through 10-fold cross validation approach. To assess the performance of each model receiver operating curve (ROC) and the corresponding area under the curve (AUC) values were computed. The model output was calibrated using Platt’s scaling and the impact of this calibration was visualized by comparing the Brier score between the uncalibrated and the calibrated outputs.

Participants were stratified into three groups based on the tertiles of the ten-year survival probability predicted by the model. The discriminative ability of the model was further validated by employing the log-rank test to compare the survival curves among these groups.

### Feature importance based on machine learning models

To estimate feature importance ranking, as well as main effect of features and interaction effect between features, SHAP (Shapley Additive explanations) was employed. The SHAP is a useful and classical method to calculate the marginal contribution of features to the model’s output. This method provides insight from both global and local perspectives, particularly beneficial for interpreting "black box model".

## Result

### Baseline characteristics

The cohort consisted of 7921 participants, with average age of 60.79±12.18, and 3866(48.81%) males. During an average follow-up period of 9.75 years, there were 1,911 deaths (24.13%), with 585 cases attributed to cardiovascular diseases. The detailed information was shown in the **Table 1**. In terms of lifestyle, there are differences between the all-cause mortality group and the cardiovascular disease mortality group and the alive group.

**Table 1.**
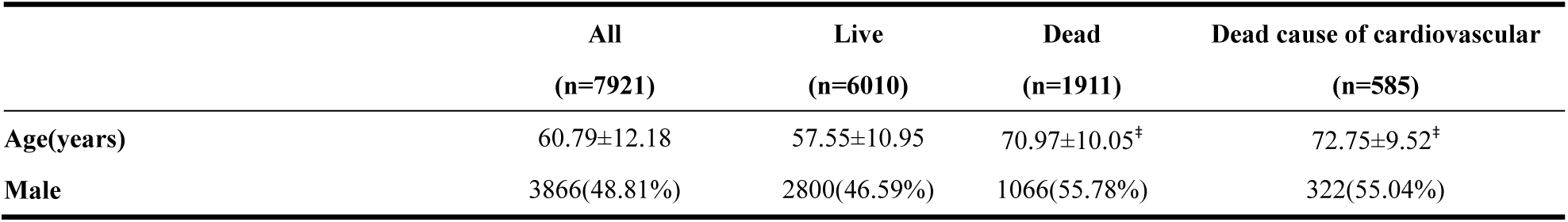
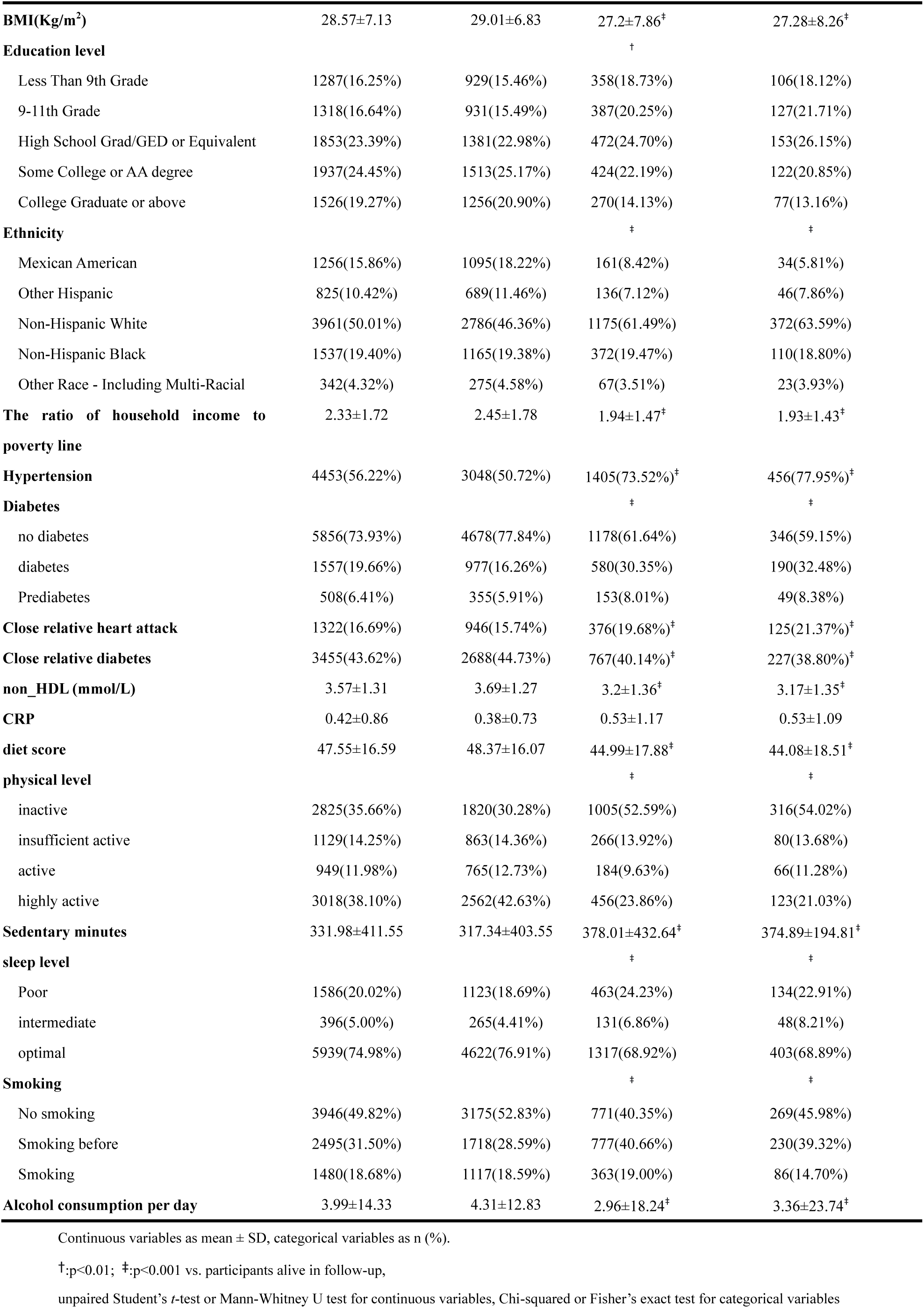
Baseline characteristics of participants.

### Performance of models

**Table 2** presents the AUC scores for all models in predicting all-cause mortality and cardiovascular disease mortality. XGBoost demonstrated notable performance, achieving an AUC score of 0.848 for predicting all-cause mortality and 0.829 for predicting cardiovascular disease mortality, establishing it as the top-performing model. The grid search parameters dictionary and the optimal parameter values were displayed in the **Supplementary Table 2**. Following calibration, there was an improvement in Brier scores, and detailed information was described in the **Supplementary Table 3**. Figure 1 shows the calibrated and uncalibrated AUC scores of the XGBoost model. The calibrated score was 0.884, indicating that the model fits the data well.

**Figure 1.**
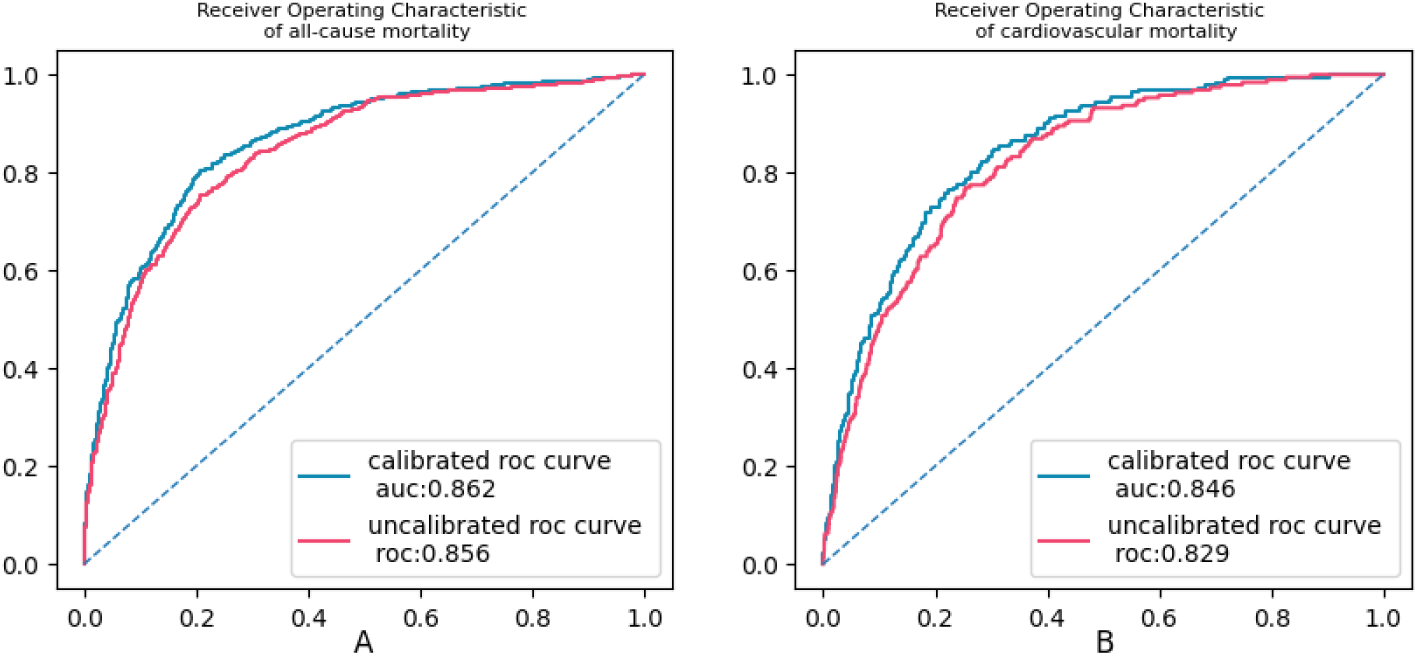
uncalibrated and calibrated ROC of XGBoost model

**Table 2.**
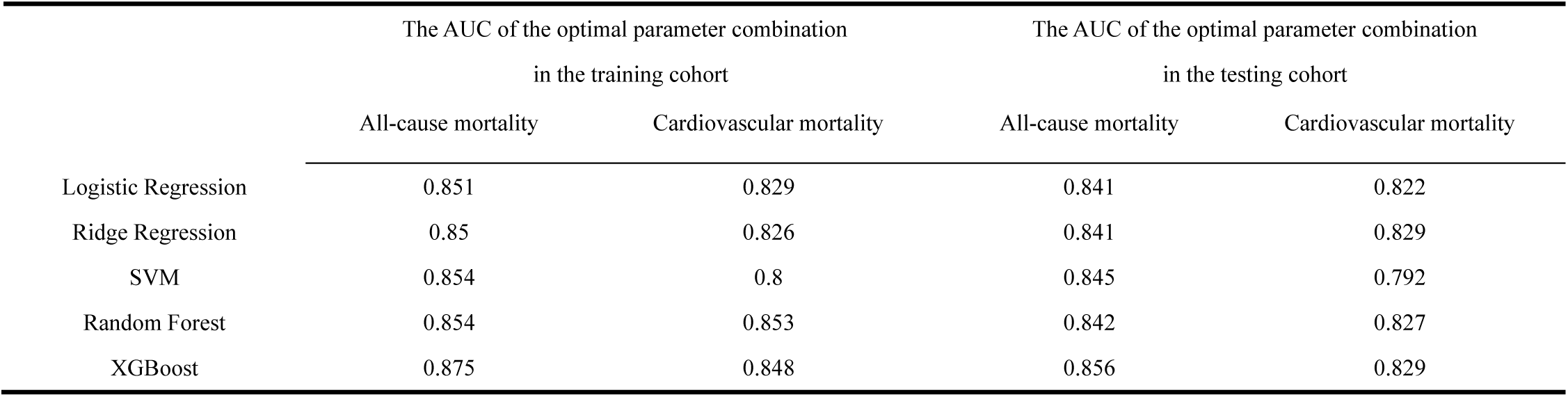
AUC score of all models.

### Machine learning-based risk stratification

Depends on the calibrated output, participants were divided into three groups. Each group survival curve was shown in **Figure 2**. It can be seen from **Supplementary Table 4** that there are significant differences in the survival curves for each group. This demonstrates that the model effectively distinguishes individuals with different risks of mortality.

**Figure 2.**
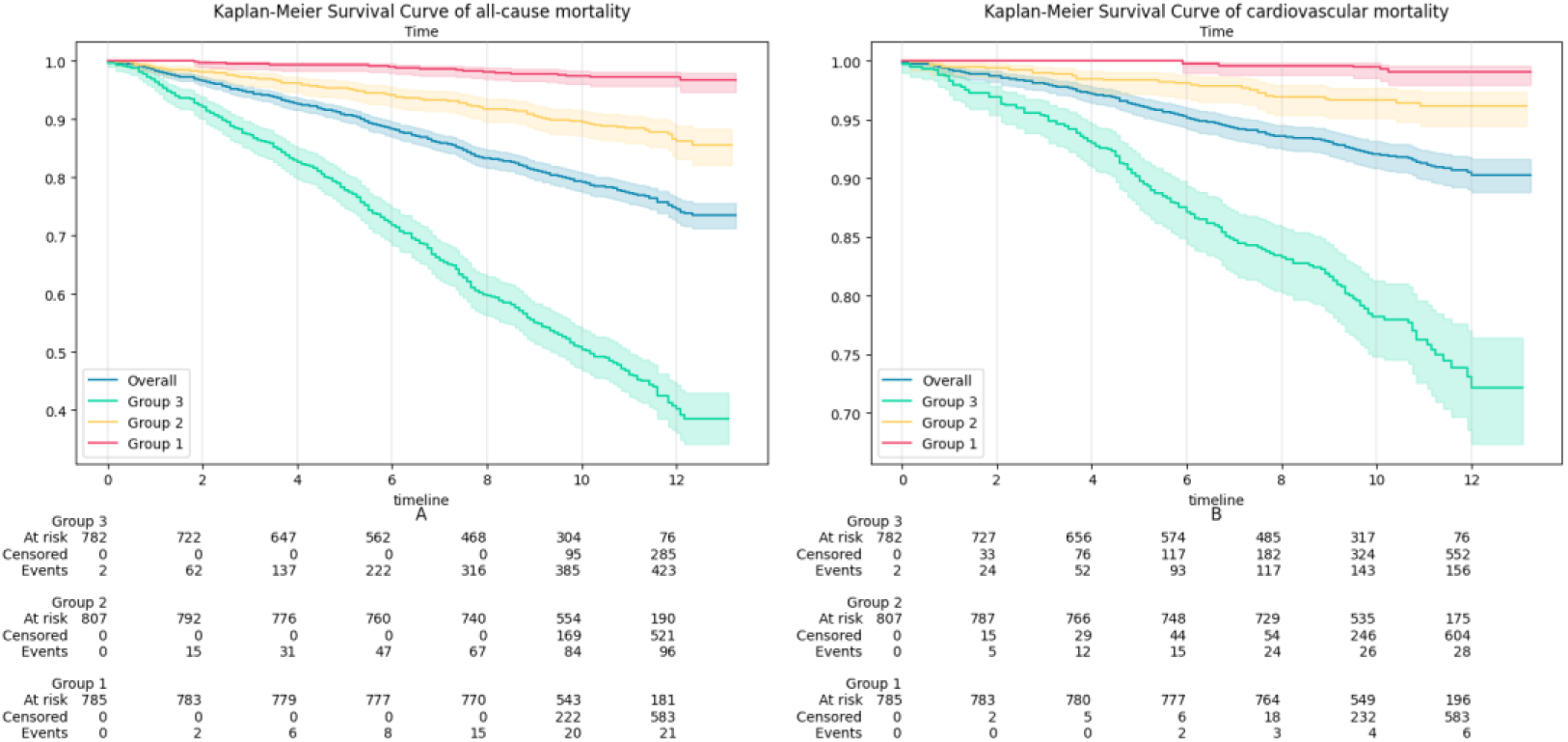
KM-curves of all groups based on tertiles

### Features importance and Features’ Role in the Model

In the prediction of both all-cause mortality and cardiovascular disease mortality, age, gender, and diabetes status have made significant contributions to the predictive outcomes (**Figure 3**). In terms of lifestyle, smoking, alcohol consumption, and physical activity emerge as significant features exerting a substantial impact on the prediction of all-cause mortality. On the other hand, the model indicates that, reduced sedentary time, higher dietary scores, and increased physical activity in the model will lower individual risk scores.

**Figure 3.**
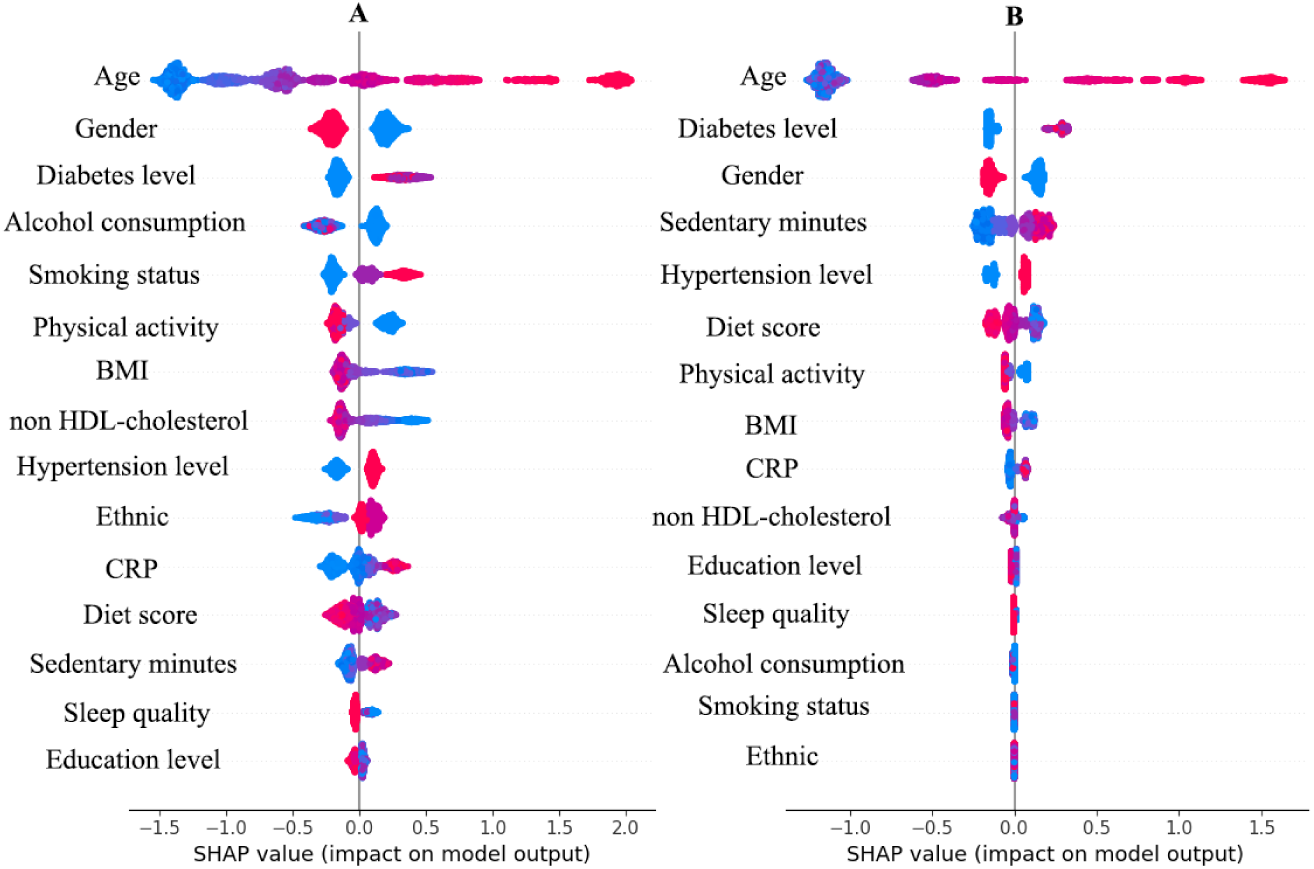
The importance and role of features in the model Figure 3 illustrates the importance of features in the model, wherein each scatter represents a sample. The importance increases from bottom to top, with color representing the numerical value of the feature. The x-axis represents the role of different values of each feature in the model, with positive values indicating that the feature increases the probability of the model making death predictions. Plot A : SHAP value of features in predicting all-cause mortality. Plot B: SHAP value of features in predicting in cardiovascular mortality.

### Features interaction effect

Given the prominent role of age in the model predictions, it is essential to further explore the interaction between age and various lifestyle factors. As shown in **Figure 4**, the impact of diet score and sedentary time on outcome prediction becomes more pronounced with advancing age, while the impact of physical activity level exhibits an opposite trend.

**Figure 4.**
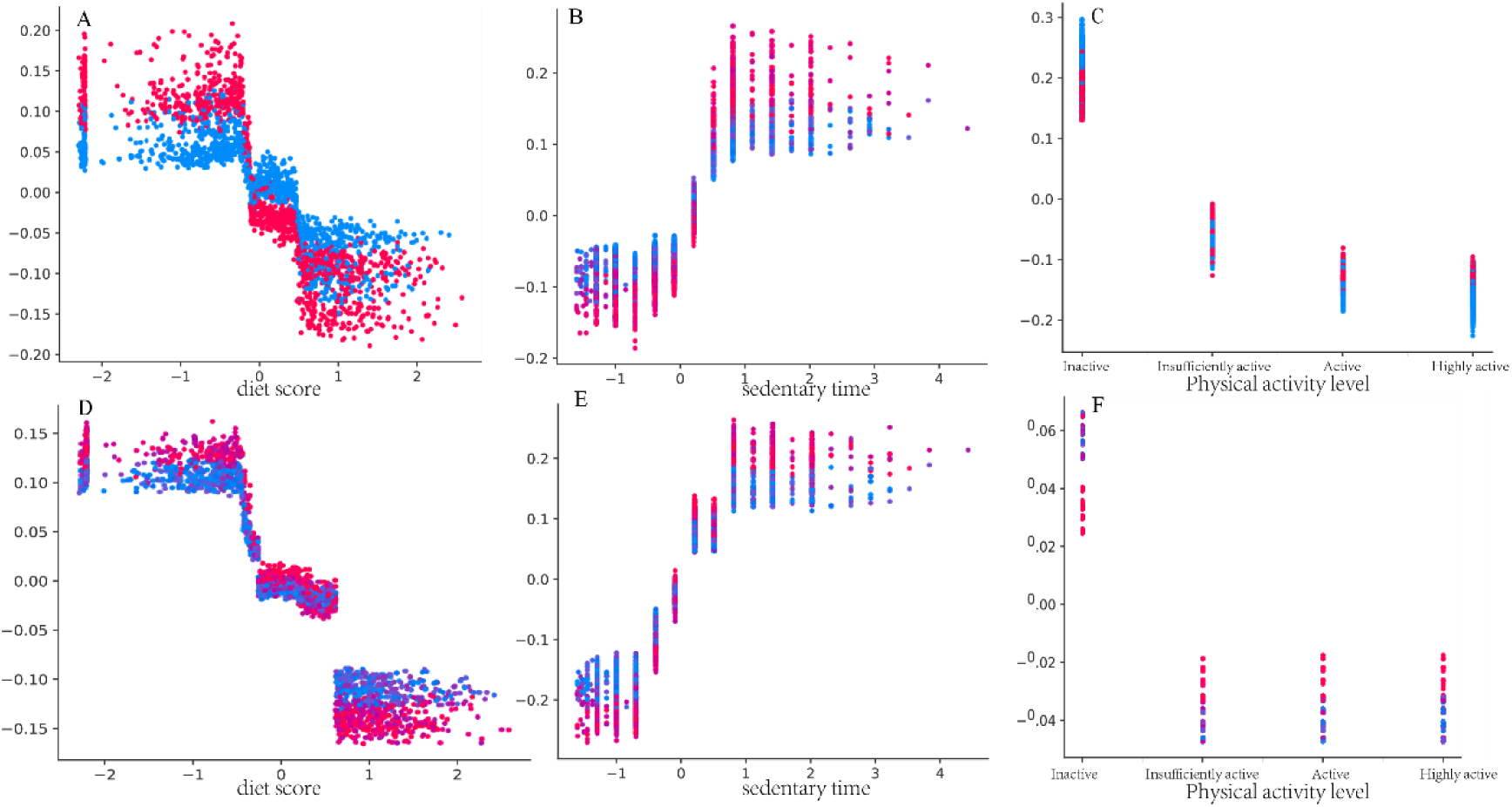
dependence plot interaction effect with age The figure illustrates the interaction between features and age. Each scatter point along a line perpendicular to the x-axis represents the SHAP values of features sharing the same x-value but varying age values. In the visualization, color represents the value of age, transitioning from blue to red as age increases. The y-axis corresponds to the SHAP values of features at different age values. A,B and C represent SHAP value of diet score, sedentary time and physical activity level in predicting all-cause mortality. D,E and F represent shap value of diet score, sedentary time and physical activity level in predicting cardiovascular mortality.

## Discussion

In this prospective cohort spanning an average of 9.75 years, a model was developed and validated to predict both the all-cause mortality and cardiovascular mortality based on the comprehensive dataset encompassing lifestyle data and basic characteristic variables. In addition, the effect of lifestyles on all-cause mortality and cardiovascular mortality and their interaction effect with age were estimated using SHAP. These estimates indicate that lifestyle affects outcome predictions to varying degrees and exhibits diverse patterns in interaction with age.

Simultaneously interpreting multiple risk factors for individual outcomes poses a challenge for the general public, as well as for healthcare professionals and policymakers. By employing ML algorithms, we established a predictive model related to lifestyle and further explored the contributions of diverse factors to survival outcomes. The results indicate that our model performs effectively and can unveil the roles of less influential predictive factors within the model. Additionally, the potential impact of complex and subtle interactions among predictive factors is often overlooked. The inherent advantages of tree models, coupled with their integration with SHAP, allowed for exploring interactions among various predictive factors.

According to the report from the Physical Activity Guidelines for Americans 2nd edition, there is a positive correlation between sedentary time and all-cause mortality^17^. A prospective survey study from NHANES reveals that, with the prolongation of sedentary time, the risk of all-cause mortality also increases^18^. Similarly, a longitudinal survey study conducted in China also identified an association between sedentary behavior and all-cause mortality^19^. In our model, sedentary behavior contributes to the model’s inclination to predict adverse events, consistent with previous research. Furthermore, we found that sedentary behavior has a stronger impact on cardiovascular mortality, ranking higher in feature importance analysis. The relationships between lifestyle factors such as physical activity^20^, diet^21^, sleep^22^, and both all-cause mortality and cardiovascular mortality have been described in detail in previous literature and is consistent with our findings. Overall, machine learning models and traditional models have drawn similar conclusions regarding the relationship between lifestyle factors and mortality.

Beyond lifestyle factors, age and gender, two fundamental demographic characteristics, play a significant role in the model. While a minority of studies may suggest that the role of age in their models is not statistically significant, the prevailing body of research, including our findings, consistently indicates that age plays a non-negligible role in outcome prediction^23,24^. Studies in various countries and regions consistently indicate that females tend to have lower mortality rates or death risks compared to males^25–27^. This finding is also reflected in our model, where the male gender feature inclines the model toward predicting a higher likelihood of death. This may be attributed to a higher proportion of females adopting healthier lifestyles compared to males^28^. Additionally, relatively higher estrogen levels in females may contribute to maintaining healthy vascular function^29^. Moreover, females might be more inclined to proactively address health issues and seek early treatment^30^.

Leveraging the advantages of tree-based models in exploring interactions in machine learning^31^, we conducted additional analysis to scrutinize the interactions between various lifestyle factors and age. We discovered some phenomena worth discussing by exploring interactions in the model through SHAP (Shapley Additive explanations).. For example, as age increases, the impact of diet and sedentary behavior on adverse outcome events gradually strengthens, while the effect of physical activity diminishes. Specifically, the gap between recommended and not recommended diet and sedentary behaviors widens across different age groups, while the gap between recommended and not recommended physical activities gradually narrows. Given the limited literature on the interaction between lifestyle and age, more research is needed to confirm this finding. The occurrence of this finding may be attributed to the insufficient granularity in the categorization of physical activity. We classified physical activity as a categorical variable with four levels. However, as participants age, although their physical activity levels decrease, they still fall within the same category as relatively younger individuals, resulting in attenuation of its impact. This can be observed in **Supplementary Table 7**; as age increases, the average exercise time in the same physical activity group gradually decreases. However, this does not imply that the role of physical activity can be disregarded in the elderly population. One reason is that low physical activity levels can exacerbate the adverse effects of sedentary behavior^32,33^.

There are countless factors associated with mortality outcomes, and it’s not practical to include all relevant variables in a predictive model. While lifestyle may not be the most significant factor in outcome prediction among many related variables, it possesses an excellent feature—modifiability. Policymakers or healthcare professionals can raise public awareness and guide individuals toward healthier lifestyles through various means such as education and outreach. Our model enables users to predict mortality based on their current conditions, serving as a warning and reminder. This functionality assists users in moving towards healthier lifestyle changes. That’s why we chose to establish a predictive model for lifestyle-related mortality rates.

### Strength and limitations

This research has several advantages and limitations that need to be acknowledged. We utilized sufficient data and implemented measures such as 10-fold cross-validation to ensure and validate the stability of the model. However, it’s important to note that our data is derived from a single cohort, and the effectiveness of the model lacks external validation. This study is a prospective cohort study, and the reliability of causal inference is relatively strong. However, during the follow-up process, a small fraction of participants were lost to follow-up (LTFU) or withdrew from the study for various reasons, leading to the possibility of not capturing the occurrence of outcome events. To the best of our knowledge, this study represents the first attempt to apply ML algorithms to explore the relationship between lifestyle and mortality. Additionally, we leveraged the advantages of tree models to investigate interactions in this context. However, inferences about the role of features based on ML only describe the features’ impact on outcome prediction within the model and may not necessarily reflect their real-world effects. The actual effects require further assessment in conjunction with domain expertise.

## Conclusion

By employing modifiable lifestyle factors and readily available indicators, we effectively predicted overall mortality and cardiovascular disease mortality using the XGBoost model. This model can serve as a valuable predictive tool to encourage individuals to modify unhealthy lifestyles and prevent adverse events.

## Data Availability

Data openly available in a public repository.

https://www.cdc.gov/nchs/nhanes/

## Reference

1. Organization WH. Call for public comments on the draft WHO Guidelines: Saturated fatty acid and trans-fatty intake for adults and children. World Health Organization. https://www.who.int/news-room/articles-detail/call-for-public-comments-on-the-draft-who-guidelines--saturated-fatty-acid-and-trans-fatty-intake-for-adults-and-children. Accessed 2018-02--06.

2. Roth GA, Mensah GA, Johnson CO, Addolorato G, Ammirati E, Baddour LM, Barengo NC, Beaton AZ, Benjamin EJ, Benziger CP, et al. Global Burden of Cardiovascular Diseases and Risk Factors, 1990-2019: Update From the GBD 2019 Study. J Am Coll Cardiol. 2020;76:2982–3021. doi: 10.1016/j.jacc.2020.11.010

3. Organization WH. WHO MORTALITY DATABASE Interactive platform visualizing mortality data. World Health Organization. https://platform.who.int/mortality/themes/theme-details/mdb/noncommunicable-diseases. Accessed 2023-12-25.

4. Khraishah H, Alahmad B, Ostergard RL, Jr., AlAshqar A, Albaghdadi M, Vellanki N, Chowdhury MM, Al-Kindi SG, Zanobetti A, Gasparrini A, et al. Climate change and cardiovascular disease: implications for global health. Nat Rev Cardiol. 2022;19:798–812. doi: 10.1038/s41569-022-00720-x

5. Li Y, Pan A, Wang DD, Liu X, Dhana K, Franco OH, Kaptoge S, Di Angelantonio E, Stampfer M, Willett WC, et al. Impact of Healthy Lifestyle Factors on Life Expectancies in the US Population. Circulation. 2018;138:345–355. doi: 10.1161/circulationaha.117.032047

6. Lu Q, Zhang Y, Geng T, Yang K, Guo K, Min X, He M, Guo H, Zhang X, Yang H, et al. Association of Lifestyle Factors and Antihypertensive Medication Use With Risk of All-Cause and Cause-Specific Mortality Among Adults With Hypertension in China. JAMA Netw Open. 2022;5:e2146118. doi: 10.1001/jamanetworkopen.2021.46118

7. Sotos-Prieto M, Bhupathiraju SN, Mattei J, Fung TT, Li Y, Pan A, Willett WC, Rimm EB, Hu FB. Association of Changes in Diet Quality with Total and Cause-Specific Mortality. N Engl J Med. 2017;377:143–153. doi: 10.1056/NEJMoa1613502

8. Kline A, Wang H, Li Y, Dennis S, Hutch M, Xu Z, Wang F, Cheng F, Luo Y. Multimodal machine learning in precision health: A scoping review. NPJ Digit Med. 2022;5:171. doi: 10.1038/s41746-022-00712-8

9. Ngiam KY, Khor IW. Big data and machine learning algorithms for health-care delivery. Lancet Oncol. 2019;20:e262–e273. doi: 10.1016/s1470-2045(19)30149-4

10. Cui F, Yue Y, Zhang Y, Zhang Z, Zhou HS. Advancing Biosensors with Machine Learning.ACS Sens. 2020;5:3346–3364. doi: 10.1021/acssensors.0c01424

11. Al’Aref SJ, Anchouche K, Singh G, Slomka PJ, Kolli KK, Kumar A, Pandey M, Maliakal G, van Rosendael AR, Beecy AN, et al. Clinical applications of machine learning in cardiovascular disease and its relevance to cardiac imaging. Eur Heart J. 2019;40:1975–1986. doi: 10.1093/eurheartj/ehy404

12. Deo RC. Machine Learning in Medicine. Circulation. 2015;132:1920–1930. doi: 10.1161/circulationaha.115.001593

13. Prevention CfDCa. About the National Health and Nutrition Examination Survey. Centers for Disease Control and Prevention. https://www.cdc.gov/nchs/nhanes/about_nhanes.htm. Accessed 2023-12-26.

14. Zhang YB, Chen C, Pan XF, Guo J, Li Y, Franco OH, Liu G, Pan A. Associations of healthy lifestyle and socioeconomic status with mortality and incident cardiovascular disease: two prospective cohort studies. Bmj. 2021;373:n604. doi: 10.1136/bmj.n604

15. Liu J, Micha R, Li Y, Mozaffarian D. Trends in Food Sources and Diet Quality Among US Children and Adults, 2003-2018. *JAMA Netw Open*. 2021;4:e215262. doi: 10.1001/jamanetworkopen.2021.5262

16. Wang C, Bangdiwala SI, Rangarajan S, Lear SA, AlHabib KF, Mohan V, Teo K, Poirier P, Tse LA, Liu Z, et al. Association of estimated sleep duration and naps with mortality and cardiovascular events: a study of 116 632 people from 21 countries. Eur Heart J. 2019;40:1620–1629. doi: 10.1093/eurheartj/ehy695

17. Promotion OoDPaH. Physical Activity Guidelines for Americans 2^nd^ edition. In; 2018:21–22.

18. Cao C, Friedenreich CM, Yang L. Association of Daily Sitting Time and Leisure-Time Physical Activity With Survival Among US Cancer Survivors. JAMA Oncol. 2022;8:395–403. doi: 10.1001/jamaoncol.2021.6590

19. Lin Y, Liu Q, Liu F, Huang K, Li J, Yang X, Wang X, Chen J, Liu X, Cao J, et al. Adverse associations of sedentary behavior with cancer incidence and all-cause mortality: A prospective cohort study. J Sport Health Sci. 2021;10:560–569. doi: 10.1016/j.jshs.2021.04.002

20. Mok A, Khaw KT, Luben R, Wareham N, Brage S. Physical activity trajectories and mortality: population based cohort study. Bmj. 2019;365:l2323. doi: 10.1136/bmj.l2323

21. Naghshi S, Sadeghi O, Willett WC, Esmaillzadeh A. Dietary intake of total, animal, and plant proteins and risk of all cause, cardiovascular, and cancer mortality: systematic review and dose-response meta-analysis of prospective cohort studies. Bmj. 2020;370:m2412. doi: 10.1136/bmj.m2412

22. Svensson T, Saito E, Svensson AK, Melander O, Orho-Melander M, Mimura M, Rahman S, Sawada N, Koh WP, Shu XO, et al. Association of Sleep Duration With All- and Major-Cause Mortality Among Adults in Japan, China, Singapore, and Korea. JAMA Netw Open. 2021;4:e2122837. doi: 10.1001/jamanetworkopen.2021.22837

23. Huang J, Liao LM, Weinstein SJ, Sinha R, Graubard BI, Albanes D. Association Between Plant and Animal Protein Intake and Overall and Cause-Specific Mortality. JAMA Intern Med. 2020;180:1173–1184. doi: 10.1001/jamainternmed.2020.2790

24. Davis JS, Banfield E, Lee HY, Peng HL, Chang S, Wood AC. Lifestyle behavior patterns and mortality among adults in the NHANES 1988-1994 population: A latent profile analysis. Prev Med. 2019;120:131–139. doi: 10.1016/j.ypmed.2019.01.012

25. Zhou T, Yuan Y, Xue Q, Li X, Wang M, Ma H, Heianza Y, Qi L. Adherence to a healthy sleep pattern is associated with lower risks of all-cause, cardiovascular and cancer-specific mortality. J Intern Med. 2022;291:64–71. doi: 10.1111/joim.13367

26. Yun JE, Won S, Kimm H, Jee SH. Effects of a combined lifestyle score on 10-year mortality in Korean men and women: a prospective cohort study. BMC Public Health. 2012;12:673. doi: 10.1186/1471-2458-12-673

27. Tamakoshi A, Tamakoshi K, Lin Y, Yagyu K, Kikuchi S. Healthy lifestyle and preventable death: findings from the Japan Collaborative Cohort (JACC) Study. Prev Med. 2009;48:486–492. doi: 10.1016/j.ypmed.2009.02.017

28. Zhu N, Yu C, Guo Y, Bian Z, Han Y, Yang L, Chen Y, Du H, Li H, Liu F, et al. Adherence to a healthy lifestyle and all-cause and cause-specific mortality in Chinese adults: a 10-year prospective study of 0.5 million people. Int J Behav Nutr Phys Act. 2019;16:98. doi: 10.1186/s12966-019-0860-z

29. Morselli E, Santos RS, Criollo A, Nelson MD, Palmer BF, Clegg DJ. The effects of oestrogens and their receptors on cardiometabolic health. Nat Rev Endocrinol. 2017;13:352–364. doi: 10.1038/nrendo.2017.12

30. Courtenay WH. Constructions of masculinity and their influence on men’s well-being: a theory of gender and health. Soc Sci Med. 2000;50:1385–1401. doi: 10.1016/s0277-9536(99)00390-1

31. Chen T, Guestrin C. Xgboost: A scalable tree boosting system. Paper/Poster presented at: Proceedings of the 22nd acm sigkdd international conference on knowledge discovery and data mining; 2016;

32. Bull FC, Al-Ansari SS, Biddle S, Borodulin K, Buman MP, Cardon G, Carty C, Chaput JP, Chastin S, Chou R, et al. World Health Organization 2020 guidelines on physical activity and sedentary behaviour. Br J Sports Med. 2020;54:1451–1462. doi: 10.1136/bjsports-2020-102955

33. Li S, Lear SA, Rangarajan S, Hu B, Yin L, Bangdiwala SI, Alhabib KF, Rosengren A, Gupta R, Mony PK, et al. Association of Sitting Time With Mortality and Cardiovascular Events in High-Income, Middle-Income, and Low-Income Countries. JAMA Cardiol. 2022;7:796–807. doi: 10.1001/jamacardio.2022.1581

